# Association of mRNA COVID-19 vaccination with overall survival in patients receiving immune checkpoint inhibitors: a retrospective cohort analysis

**DOI:** 10.64898/2026.01.19.26344403

**Authors:** Mark TK Cheng, Jodi L Keen, Andrew J Dooley, Lesley Arends, Steven Frost, David M Favara

**Affiliations:** Department of Haematology, Cambridge University Hospitals NHS Foundation Trust, Cambridge, UK; Cambridge Institute for Therapeutic Immunology and Infectious Disease, University of Cambridge, Cambridge, UK; Department of Information Management, Cambridge University Hospitals NHS Foundation Trust, Cambridge, UK; Department of Oncology, University of Cambridge, Cambridge, UK; Department of Oncology, Cambridge University Hospitals NHS Foundation Trust, Cambridge, UK; Cancer Directorate, Cambridge University Hospitals NHS Foundation Trust, Cambridge, UK; Department of Structural Studies, MRC Laboratory of Molecular Biology, Cambridge, UK

**Keywords:** immune checkpoint inhibitors, COVID-19 vaccination, mRNA vaccines, cancer immunotherapy, overall survival, SARS-CoV-2

## Abstract

**Background:** A retrospective study in Nature recently reported improved overall survival (OS) when COVID-19 mRNA vaccination was administered around initiation of immune checkpoint inhibitor (ICI) therapy. Whether this association depends on vaccination timing relative to ICI treatment remains unclear.

**Methods:** We conducted a single-centre retrospective cohort study of patients receiving palliative-intent ICI therapy at a UK tertiary cancer centre between October 2014 and December 2025. Two vaccination exposure definitions were evaluated: (1) vaccination from 100 days before to 100 days after the first ICI cycle (initial window); and (2) vaccination from 100 days before the first ICI cycle to 100 days after the final ICI cycle (extended window). Multivariable Cox models adjusted for age, sex, treatment line and year of ICI initiation, with the baseline hazard stratified by tumour group. Landmark, time-varying, pre-ICI, active vaccinated comparator and calendar-restricted analyses were performed.

**Results:** Among 2,054 patients, 493 (24.0%) received ≥1 COVID-19 vaccination. In the initial fixed-window analysis, mRNA vaccination was not significantly associated with OS (HR 0.80; 95% CI 0.60-1.06; *p*=0.123). No significant association was observed in day-100 landmark, initial time-varying or pre-ICI analyses. The active vaccinated comparator analysis was also non-significant. In the extended fixed-window analysis, longer OS was associated with mRNA (HR 0.59; 95% CI 0.46-0.76; *p*<0.001) and adenoviral-vector vaccination (HR 0.65; 95% CI 0.51-0.83; *p*<0.001). The mRNA association was attenuated in calendar-restricted analyses and was non-significant in the supportive time-varying analysis (HR 0.82; 95% CI 0.66-1.04; *p*=0.099).

**Conclusions:** Survival associations with COVID-19 mRNA vaccination varied according to exposure definition and analytic handling of vaccination timing. The strong association observed in the extended fixed-window analysis was attenuated in calendar-restricted and supportive time-varying analyses. Prospective studies are required to determine whether COVID-19 mRNA vaccination at specific time points during ICI therapy influences clinical outcomes.

**Highlights:**

1. Initial fixed-window mRNA vaccination was not significantly associated with overall survival.
2. Extended fixed-window analyses showed survival associations that were attenuated in time-varying analyses.
3. Landmark, pre-ICI and active vaccinated comparator analyses were non-significant.
4. Prospective studies are required to assess causality and tumour-specific effects.

## Introduction

Grippin *et al*. recently reported a strong retrospective association between COVID-19 mRNA vaccination and improved overall survival (OS) among American patients with advanced non-small cell lung cancer (NSCLC) and metastatic melanoma receiving immune checkpoint inhibitor (ICI) therapy. Patients who received an mRNA vaccine within 100 days before or after ICI initiation experienced significantly improved OS compared with unvaccinated patients (Grippin et al., 2025). This association was not observed for influenza or pneumococcal protein vaccination in NSCLC cohorts (Grippin et al., 2025), suggesting a potential mRNA vaccine-specific effect. Supporting preclinical results showed that non-tumour antigen mRNA vaccines can induce a type I interferon-mediated innate immune response, associated with increased tumour PD-L1 expression and enhanced antigen-presenting cell activation, potentially augmenting tumour-specific T cell responses and sensitivity to ICI therapy (Grippin et al., 2025).

Given the potential clinical implications of these findings, independent validation in different real-world populations is warranted. In particular, it remains unclear whether the reported survival association is confined to vaccination administered at or around the time of ICI initiation, or whether vaccination occurring across a broader ICI treatment period may also be associated with improved outcomes. We therefore retrospectively evaluated the association between COVID-19 vaccination timing and OS in a single-centre United Kingdom (UK) cohort of patients treated with palliative-intent ICI therapy across multiple tumour types.

## Methods

### Study design and cohort

A retrospective cohort analysis was conducted using the electronic patient record (EPR) system at Addenbrooke’s Hospital, Cambridge University Hospitals NHS Foundation Trust, Cambridge, UK. Ethical approval was granted by Cambridge University Hospitals NHS Foundation Trust (ID7592; PRN13592). The EPR was queried to identify all patients treated with UK-licensed immune checkpoint inhibitor (ICI) therapy (atezolizumab, avelumab, cemiplimab, dostarlimab, durvalumab, ipilimumab, nivolumab, pembrolizumab, relatlimab, and tremelimumab) between 1 October 2014 and 19 December 2025.

Demographic characteristics, cancer diagnosis information, ICI treatment details (indication, treatment intent, line of therapy, start/end dates, number of ICI cycles), survival data (date of death or last follow-up) and COVID-19 vaccination records (vaccination status, vaccine type, and dates of administration) were extracted from the EPR. COVID-19 vaccination records within the EPR included hospital vaccination records and information from regional GP practices, collated by the hospital as part of the regional COVID-19 programme. Patients alive at the time of database extraction were censored on 19 December 2025. Where discrepancies or ambiguities were identified, patient records were manually reviewed to ensure data accuracy.

As tumour stage was not consistently available within the EPR, treatment intent recorded at time of ICI initiation was used as a pragmatic surrogate for disease extent. Patients were therefore categorised into two groups: curative-intent (comprising neoadjuvant and adjuvant ICI therapies) and palliative-intent (comprising non-curative ICI therapy for advanced disease, including metastatic or unresectable disease). Where intent records were missing or ambiguous, individual patient records were manually reviewed to determine whether treatment was delivered with curative or palliative intent. The validity of palliative-intent treatment as a surrogate for advanced disease was examined in patients with available stage or resectability information. To minimise baseline disease-stage imbalances between vaccination exposure groups, all survival analyses were restricted to patients treated with palliative-intent ICI therapy.

### Vaccination exposure definitions

To evaluate the association between COVID-19 vaccination and survival, two vaccination exposure definitions were examined. The initial exposure window mirrored that of Grippin *et al*. (Grippin et al., 2025), classifying patients as vaccinated if they received at least one COVID-19 vaccine dose from 100 days before to 100 days after the first palliative-intent ICI cycle. The extended exposure window classified patients as vaccinated if they received at least one COVID-19 vaccine dose from 100 days before the first palliative-intent ICI cycle (C1) to 100 days after the final ICI cycle. Vaccinations were classified by platform as mRNA, adenoviral-vector (AdV) or mixed. In the conventional fixed-window analyses, patients with no recorded COVID-19 vaccination at any time formed the unvaccinated comparator group, while patients vaccinated exclusively outside the relevant window were excluded from that comparison. mRNA and AdV vaccination were analysed separately using the same methods. Patients who received both mRNA and AdV vaccines within the relevant exposure window were classified as mixed and excluded from the platform-specific fixed-window comparisons.

### Outcome, covariates and statistical analyses

Summary statistics were calculated for all palliative-intent treated patients, stratified by vaccine exposure and tumour type. Associations between vaccination exposure and overall survival (OS) were assessed using Cox proportional hazards models (Cox, 1972). The primary outcome was OS defined as the time from the first palliative-intent ICI cycle to death from any cause. Patients without a recorded death were censored on 19 December 2025. Follow-up was truncated at 60 months.

The core adjustment set comprised age, sex, line of systemic therapy at palliative-intent ICI initiation and calendar year of ICI initiation, with the baseline hazard stratified by tumour group (kidney cancer; melanoma; non-small cell lung cancer (NSCLC); or remaining tumour types). Year of ICI initiation was modelled continuously in the core model and categorically in a sensitivity analysis. Pre-ICI performance status was standardised as ECOG 0, 1 or ≥2 and incorporated in additional sensitivity analyses. As comorbidity data were incomplete, recorded non-cancer comorbidity burden was evaluated in available-case sensitivity analyses rather than included in the core model. Among patients with available non-cancer comorbidity data, the number of recorded comorbidities was included as a categorical covariate. BMI was not included as an adjustment variable because pre-treatment measurements were not widely available.

Three analyses were given greatest weight in assessing time-related bias. For the day-100 landmark analysis, patients were required to be alive and under observation beyond 100 days from their first palliative-intent ICI cycle. Follow-up began on day 100 and vaccination exposure was defined using the initial -100 to +100-day window. Patients vaccinated before the landmark but exclusively outside this window were excluded from the pairwise comparison, while vaccination occurring after day 100 did not alter landmark classification.

In a time-varying analysis of the initial exposure window, patients vaccinated during the 100 days before starting ICI treatment were considered vaccinated from the start of follow-up. Patients vaccinated during the first 100 days after starting ICI treatment contributed follow-up times to the unvaccinated group until the date of vaccination and to the vaccinated group thereafter. mRNA and AdV vaccination were modelled simultaneously with standard errors accounting for repeated follow-up intervals within individual patients. The time-varying analysis was also repeated using the extended exposure definition as a supportive sensitivity analysis. A separate pre-ICI analysis classified vaccination status using only vaccination history available when ICI began. Recent vaccination was defined as vaccination from 100 days before to the day of the first palliative-intent ICI cycle. Patients vaccinated before ICI initiation but with no vaccination during the preceding 100 days formed a separate previous-vaccination group, while vaccination occurring after ICI did not change the patient’s baseline classification.

To examine whether observed associations might reflect characteristics associated with receiving vaccination rather than vaccination timing itself, patients who received mRNA vaccination within 100 days before ICI initiation were compared with patients who had previously received COVID-19 vaccine but had received no vaccine during the preceding 100 days. This comparison was also repeated in patients initiating ICI therapy during 2021-2022. To assess calendar effects more directly, the fixed initial and extended analyses were also repeated in patients initiating ICI therapy during 2021-2022. Additional sensitivity analyses restricted the initial time-varying, pre-ICI and extended supportive time-varying analyses to patients initiating ICI therapy in 2021 or later. The principal vaccination exposure definitions and complementary analytical approaches are summarised schematically in **Supplementary Fig. 1**.

**Fig. 1.**
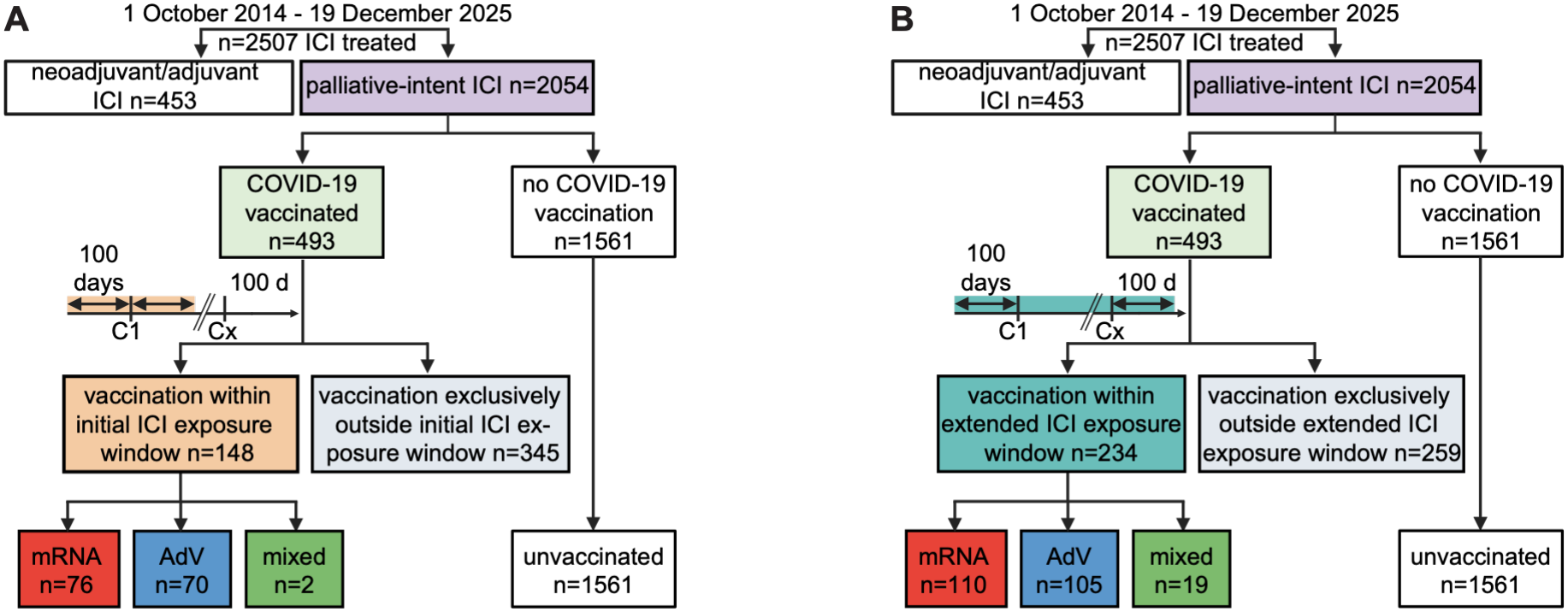
Cohort selection and COVID-19 vaccination exposure definitions. **A**, Initial exposure window, defined as receipt of at least one COVID-19 vaccine dose from 100 days before to 100 days after the first palliative-intent ICI cycle (C1). **B**, Extended exposure window, defined as receipt of at least one COVID-19 vaccine dose from 100 days before the first palliative-intent ICI cycle (C1) to 100 days after the final ICI cycle (Cx). Patients receiving neoadjuvant or adjuvant ICI therapy were excluded from survival analyses. mRNA and AdV groups indicate exposure only to that vaccine platform within the relevant window. The mixed group indicates exposure to both mRNA and AdV vaccines. Abbreviations: AdV - adenoviral-vector; ICI - immune checkpoint inhibitor.

An additional sensitivity analysis examined a shorter 30-day pre-ICI vaccination window (days -30 to 0), excluding post-ICI vaccination from exposure classification. Patients were grouped separately for mRNA and AdV vaccination as vaccinated within 30 days before ICI initiation, vaccinated more than 30 days before ICI initiation, or having no recorded pre-ICI vaccination. OS associations were assessed using the core-adjusted Cox model.

Proportional-hazards assumptions were assessed using Schoenfeld residual tests. Tumour-specific analyses were exploratory and were not adjusted for multiple comparisons. For tumour-specific analyses, line of systemic therapy was categorised as first-line versus second-line or later. Statistical significance was assessed using two-sided *p* values. All analyses were performed using R (v4.4.1) within RStudio (v2025.05.0+496). Cox proportional hazards analyses, including landmark, time-varying and sensitivity analyses, were conducted using the survival package (v3.8.3) (Therneau and Grambsch, 2000).

## Results

Between 1 October 2014 and 19 December 2025, 2,507 patients received ICI therapy at Addenbrooke’s Hospital. Of these, 453 (18.1%) received neoadjuvant or adjuvant ICI therapy whilst 2,054 (81.9%) received palliative-intent therapy for advanced disease and comprised the survival-analysis cohort (**Fig. 1a**).

Within the palliative-intent cohort, tumour types included melanoma (n=465; 22.6%), kidney cancer (n=462; 22.5%), NSCLC (n=311; 15.1%) and remaining tumour types (n=816; 39.7%) (**Supplementary Tables 1 and 3**). ICI therapy was given as first-line systemic therapy in 1,497 patients (72.9%), second-line in 382 (18.6%), third-line in 132 (6.4%) and fourth-line or later in 43 (2.1%). Among the palliative-intent ICI-treated cohort, 493 (24.0%) had at least one recorded COVID-19 vaccination, while 1,561 (76.0%) had no recorded COVID-19 vaccination. Of 1,144 recorded COVID-19 vaccine doses, 55.6% were mRNA-based (49.6% Pfizer-BioNTech BNT162b2; 6.0% Moderna mRNA-1273), and 44.4% were adenoviral-vector vaccines (AstraZeneca ChAdOx1-S).

**Table 1.**
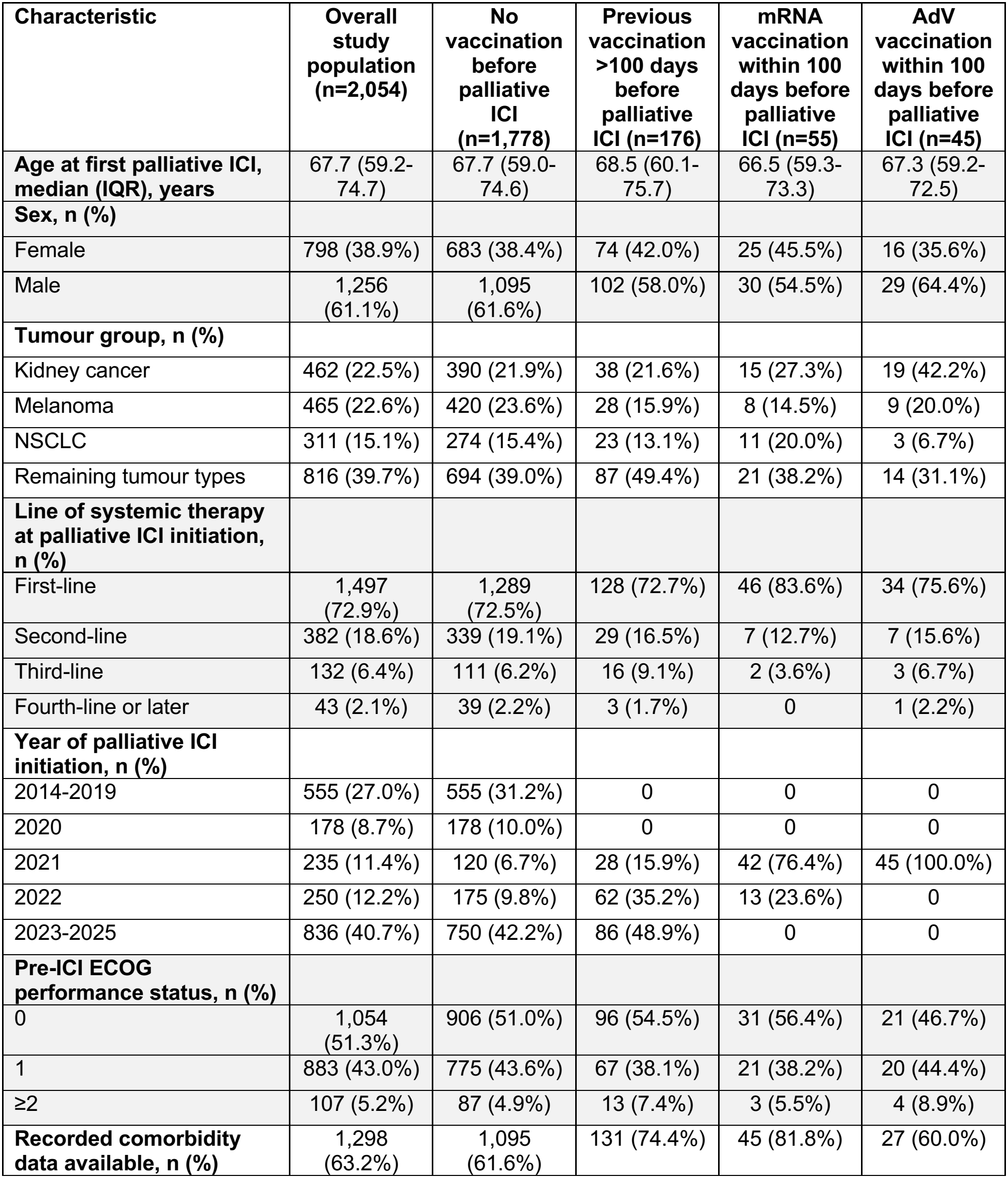
Baseline characteristics of patients receiving palliative-intent ICI therapy according to COVID-19 vaccination status at ICI initiation. Values are reported as median (IQR) or n (%) unless otherwise stated. The ‘previous vaccination >100 days before palliative ICI’ group comprised patients who had previously received a COVID-19 vaccine but had not received a vaccine within 100 days before starting palliative ICI. Pre-ICI ECOG performance status was available for 2,044 of 2,054 patients (99.5%). Abbreviations: AdV - adenoviral-vector; ECOG - Eastern Cooperative Oncology Group; ICI - immune checkpoint inhibitor; IQR - interquartile range; NSCLC - non-small cell lung cancer.

The validity of palliative treatment intent as a surrogate for advanced disease was assessed by manual EPR review of 809/2,054 (39.4%) patients treated with palliative-intent ICI therapy. At the time of ICI initiation, all 809 had metastatic or unresectable disease, supporting the use of treatment intent as a pragmatic surrogate for advanced disease. Baseline characteristics according to vaccination status at ICI initiation are shown in **Table 1**.

### Initial exposure window

Using the initial exposure window, defined to mirror that used by Grippin *et al*., 148 of 493 (30.0%) vaccinated patients received at least one COVID-19 vaccine dose from 100 days before to 100 days after their first palliative-intent ICI cycle, while 345 were vaccinated exclusively outside this window. Among vaccinated patients within this window, 76 (51.4%) received mRNA vaccines only, 70 (47.3%) received AdV vaccines only, and 2 (1.4%) had exposure to both mRNA and AdV vaccines (**Fig. 1a, Supplementary Table 1**).

In the conventional fixed-window analysis, mRNA vaccination was not significantly associated with OS after adjustment for age, sex, line of systemic therapy and year of ICI initiation, with the baseline hazard stratified by tumour group (HR 0.80; 95% CI 0.60-1.06; *p*=0.123). The corresponding estimate for AdV vaccination was HR 0.89 (95% CI 0.67-1.19; *p*=0.438) (**Table 2**).

**Table 2.**
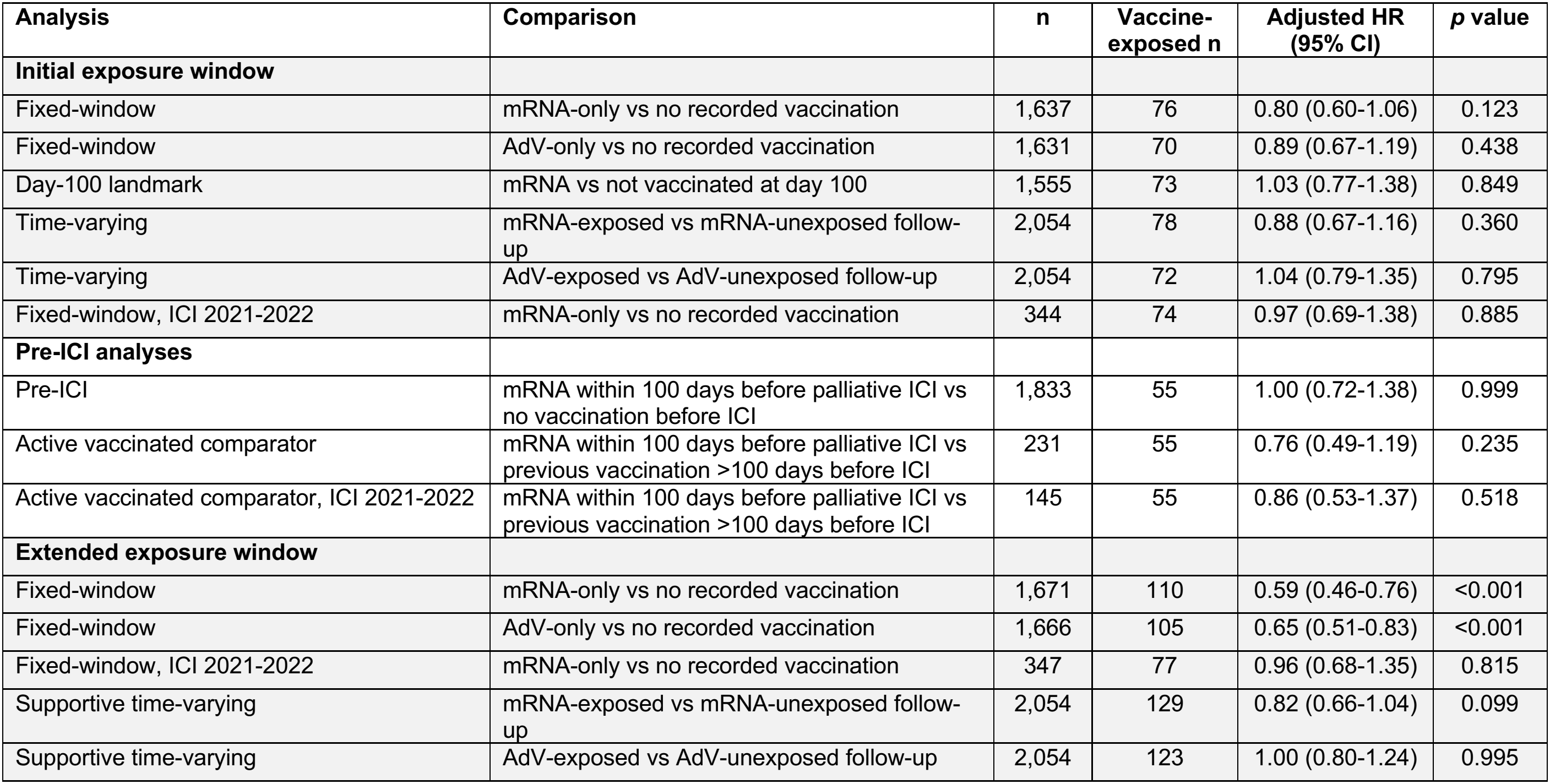
Adjusted associations between COVID-19 vaccination and overall survival in patients receiving palliative-intent ICI therapy. Hazard ratios were adjusted for age, sex, line of systemic therapy at palliative ICI initiation and year of ICI initiation, with the baseline hazard stratified by tumour group. For time-varying analyses, mRNA and AdV vaccination were modelled simultaneously. ‘Vaccine-exposed n’ indicates the number of patients in the relevant vaccinated group. In time-varying analyses, it indicates the number of patients who contributed any follow-up while classified as exposed to that vaccine type. The extended-window time-varying analysis was considered supportive because whether a vaccination qualified for the extended window could only be determined retrospectively, once the patient’s final ICI cycle was known. Follow-up was truncated at 60 months. Of 1,767 patients eligible for the day-100 landmark analysis, 212 were excluded from the mRNA comparison because of vaccination outside the exposure window (n=143), AdV-only vaccination (n=67) or mixed-platform vaccination (n=2). Abbreviations: AdV - adenoviral-vector; CI - confidence interval; HR - hazard ratio; ICI - immune checkpoint inhibitor.

Results were similar in analyses designed to reduce time-related bias. Among 1,767 patients alive and under observation beyond day 100, the day-100 landmark analysis showed no association between mRNA vaccination and subsequent OS (HR 1.03; 95% CI 0.77-1.38; *p*=0.849). In the time-varying analysis of the initial window, mRNA vaccination was also not associated with OS (HR 0.88; 95% CI 0.67-1.16; *p*=0.360), with a corresponding AdV estimate of HR 1.04 (95% CI 0.79-1.35; *p*=0.795) (**Table 2**).

When vaccination status was defined using only information available at ICI initiation, 55 patients had received an mRNA vaccine within the preceding 100 days, 45 had received an AdV vaccine, 176 had been vaccinated previously but not within the preceding 100 days, and 1,778 had not previously been vaccinated. Recent pre-ICI mRNA vaccination was not associated with OS compared with patients unvaccinated at ICI initiation (HR 1.00; 95% CI 0.72-1.38; *p*=0.999). When patients with recent pre-ICI mRNA vaccination were instead compared with previously vaccinated patients, the HR was 0.76 (95% CI 0.49-1.19; *p*=0.235). Restricting this comparison to patients initiating ICI during 2021-2022 gave a similar result (HR 0.86; 95% CI 0.53-1.37; *p*=0.518) (**Table 2**).

Calendar-restricted conventional analyses also showed no significant association. Among patients initiating ICI during 2021-2022, the adjusted HR for initial-window mRNA vaccination was 0.97 (95% CI 0.69-1.38; *p*=0.885) (**Table 2**).

To assess whether a shorter pre-ICI vaccination period influenced survival associations, analyses were performed using a 30-day window before ICI initiation (days -30 to 0; post-ICI vaccination excluded from exposure classification). mRNA vaccination within this window (n=26) was not significantly associated with OS when compared with no recorded pre-ICI vaccination (HR 0.89, 95% CI 0.55-1.44; *p*=0.637). Findings were similar when restricted to patients initiating ICI during 2021-2022 (**Supplementary Table 2**).

### Extended exposure window

Using the extended exposure window, 234 of 493 vaccinated patients (47.5%) received at least one COVID-19 vaccine dose from 100 days before the first palliative-intent ICI cycle to 100 days after the final ICI cycle, while 259 (52.5%) were vaccinated exclusively outside this window. Of those vaccinated within the extended window, 110 (47.0%) received mRNA vaccines only, 105 (44.9%) received AdV vaccines only, and 19 (8.1%) had exposure to both mRNA and AdV vaccines (**Fig. 1b**; **Supplementary Table 3**).

In the conventional adjusted fixed-window analysis, mRNA vaccination was associated with longer OS (HR 0.59; 95% CI 0.46-0.76; *p*<0.001). AdV vaccination showed a similar association (HR 0.65; 95% CI 0.51-0.83; *p*<0.001) (**Table 2**). In a calendar-restricted analysis of patients initiating ICI during 2021-2022, the adjusted HR for mRNA vaccination was 0.96 (95% CI 0.68-1.35; *p*=0.815) (**Table 2**).

In the supportive time-varying analysis using the extended exposure definition, the mRNA estimate was not statistically significant (HR 0.82; 95% CI 0.66-1.04; *p*=0.099). The corresponding time-varying AdV estimate was HR 1.00 (95% CI 0.80-1.24; *p*=0.995) (**Table 2**). Additional sensitivity analyses modelling year of ICI initiation categorically and restricting analyses by calendar era are reported in **Supplementary Table 4**.

Tumour-specific analyses were exploratory. No tumour-specific association reached statistical significance in adjusted day-100 landmark, initial time-varying, pre-ICI or extended supportive time-varying analyses. Confidence intervals were wide due to the small numbers of mRNA vaccine-exposed patients within individual tumour groups (**Supplementary Table 5**).

Sensitivity analyses incorporating pre-ICI ECOG performance status and recorded non-cancer comorbidity burden did not alter the landmark, time-varying or pre-ICI estimates. Non-cancer comorbidity data were available for 1,298 patients (63.2%) (**Supplementary Table 6**). Proportional-hazards diagnostics are reported in **Supplementary Table 7**.

## Discussion

In this single-centre UK retrospective cohort of 2,054 patients receiving palliative-intent immune checkpoint inhibitor (ICI) therapy for advanced disease across a range of tumour types, we found no robust evidence that COVID-19 mRNA vaccination was associated with improved overall survival. We first investigated two fixed exposure windows: an initial window defined as receipt of vaccination from 100 days before to 100 days after the first palliative-intent ICI cycle (corresponding to the original criteria used by Grippin *et* al.), and an extended window from 100 days before the first ICI cycle to 100 days after the final ICI cycle. Neither mRNA nor AdV vaccination was significantly associated with longer overall survival in the adjusted initial fixed-window analysis, and no significant association was observed in the day-100 landmark, initial time-varying, pre-ICI, active vaccinated comparator or calendar-restricted analyses. In contrast, the fixed-window analysis using the extended exposure window showed associations with longer overall survival for both mRNA and AdV vaccination. The mRNA association persisted when year of ICI initiation was modelled categorically but became non-significant in calendar-restricted and supportive time-varying analyses.

The divergence between fixed-window and time-varying estimates is consistent with the potential for time-related bias, including immortal-time and survivor bias, since patients vaccinated after ICI initiation must remain alive long enough to receive a qualifying vaccination. This concern is greater for the extended window, for which exposure classification also depends on the retrospectively determined final ICI cycle. To examine these potential sources of bias and confounding, year of ICI initiation was included in the core model and assessed using both continuous and categorical specifications, with additional analyses restricted by calendar era. The principal landmark, time-varying and pre-ICI estimates were not materially altered by additional adjustment for pre-ICI ECOG performance status or recorded comorbidity burden. We also compared patients receiving mRNA vaccination within 100 days before ICI initiation with patients who had been vaccinated previously but not during the preceding 100 days. This active vaccinated comparator was intended to reduce differences associated with vaccine uptake and healthcare engagement that may distinguish vaccinated from unvaccinated patients. No significant survival association was observed in this comparison, including when restricted to patients initiating ICI during 2021-2022.

Our findings should be considered alongside an evolving literature of retrospective observational studies in which both favourable and null survival associations have been reported. Grippin *et al*. first reported improved overall survival among patients with advanced melanoma and NSCLC who received mRNA COVID-19 vaccination within 100 days before or after ICI initiation, together with preclinical evidence showing that innate immune activation driven by the mRNA lipid nanoparticle platform (rather than antigen specificity) could enhance sensitivity to immune checkpoint blockade (Grippin et al., 2025). Heudel *et al*. reported improved overall survival in a retrospective pan-cancer cohort of 281 patients (80.7% metastatic) receiving mRNA vaccination within 100 days of ICI initiation (Heudel et al., 2026), while Arbel *et al*. reported a similar association among 4,407 patients with NSCLC or melanoma (disease stage unreported; the cohort included both adjuvant and palliative-intent treatment) (Arbel et al., 2026). More recently, Jourdain *et al*. reported a modest survival association in a large retrospective French cohort of 30,898 matched anti-PD-1/PD-L1 treatment initiators (63.5% metastatic; the cohort included both adjuvant and advanced disease indications) (Jourdain et al., 2026). Using a vaccine-era cohort, calendar-quarter matching, a 90-day landmark and propensity-score weighting, peri-ICI mRNA vaccination was associated with a modestly lower all-cause mortality (Jourdain et al., 2026). Our confidence intervals do not exclude a similarly modest survival association.

Other analyses have raised important questions about whether these associations represent an mRNA-specific interaction with ICI therapy. A target-trial reanalysis of the Grippin data by Dumas *et al*. found that the apparent survival benefit was substantially attenuated after aligning time zero at first ICI treatment, excluding historical controls treated before COVID-19 vaccine availability and accounting for the 100-day vaccination assignment period (Dumas et al., 2026). Similarly, a retrospective target-trial-emulation-informed TriNetX Global Collaborative Network study by Lin *et al*. used a -100 to +100-day peri-ICI vaccination window, with follow-up beginning at a prespecified day-100 landmark (Lin et al., 2026). Among 4,572 propensity-score-matched patients with lung cancer (disease stage and treatment intent not reported), peri-ICI vaccination was not associated with overall survival (Lin et al., 2026). In an independent retrospective pan-cancer cohort of 8,368 ICI-treated patients (44.6% metastatic), Jee *et al*. found that the apparent survival advantage diminished after 2021, extended to patients receiving non-ICI therapies and disappeared in analyses restricted to vaccine-eligible periods and in landmark analyses (Jee et al., 2026). In a Medicare analysis of 121,676 patients initiating anticancer therapy (which included 10,824 patients initiating ICI therapy; individual stage and tumour type distributions not reported), Karadakic *et al*. found lower mortality associated with mRNA COVID-19, adenoviral COVID-19 and influenza vaccines given within 100 days of initiating both ICI and non-ICI anticancer therapies (Karadakic et al., 2026). These associations argue against a uniquely mRNA-mediated enhancement of ICI efficacy (Jee et al., 2026, Karadakic et al., 2026).

Most recently, Salem *et al*. retrospectively analysed 95,015 patients (63.7% metastatic) initiating ICI therapy using the French health data system (Salem et al., 2026). Pre-ICI mRNA vaccination within 100 days was associated with modestly lower early hazard of all-cause mortality at 1 and 3 months, with attenuation at 6 months, but no significant association from 12 months onwards. Similar early patterns were observed with non-mRNA COVID-19 and other vaccines. The authors suggested that these findings were more consistent with a modest, transient association that was not mRNA-specific, potentially reflecting healthy-vaccinee or healthcare-engagement effects rather than providing evidence for a durable mRNA-ICI interaction (Salem et al., 2026).

Absalyamov *et al*. recently reported an association between mRNA vaccination within 30 days before ICI initiation and improved OS in a retrospective cohort of 2,888 patients (75.2% metastatic) (Absalyamov et al., 2026). Unlike studies using broader peri-ICI exposure windows, they compared vaccination within 30 days before ICI, more than 30 days before ICI, and no prior vaccination. Applying this narrower pre-ICI window to our cohort did not identify any statistically significant associations between vaccination within 30 days before ICI initiation and OS, although the small number of patients vaccinated within 30 days before ICI initiation limits interpretation.

Our study has several limitations. Its retrospective observational design precludes causal inference and residual confounding cannot be excluded despite multivariable adjustment and sensitivity analyses. The study was conducted at a single UK tertiary centre, which may limit its generalisability. The relatively small number of vaccine-exposed patients also limited statistical precision. ICI treatment spanned 2014-2025, covering both pre-vaccine and vaccine eras and substantial changes in ICI indications, treatment sequencing and supportive care. Year of ICI initiation was included in adjusted models and calendar-restricted analyses were performed, but residual calendar-era confounding may remain. COVID-19 vaccination status relied on vaccination records available within the EPR, which included data from both hospital and regional primary care services. The proportion of patients with any recorded COVID-19 vaccination initiating ICI therapy in 2021, 2022, 2023, 2024 and 2025, was 58.7%, 30.8%, 14.0%, 9.9% and 7.6%, respectively. As the completeness of vaccination capture could not be independently verified, some vaccinated patients may have been misclassified as unvaccinated. Such misclassification may have biased the estimated associations, although the direction and magnitude of this bias cannot be determined. As death ascertainment relied on hospital EPR records without independent national mortality linkage, incomplete capture of deaths cannot be excluded. Detailed disease burden, metastatic sites and PD-L1 status were not sufficiently complete for adjustment. Line of systemic therapy was available for all patients and included in the models, but detailed information on prior treatment regimens and cumulative treatment exposure was not systematically extracted for the full cohort. Landmark and time-varying analyses reduced time-related bias but could not eliminate selection effects or other sources of bias. The extended-window time-varying analysis remained limited because whether a vaccination qualified for the exposure window depended on the retrospectively determined final ICI cycle. As tumour stage was not consistently available, recorded treatment intent was used as a pragmatic surrogate for advanced disease. However, manual EPR review of 809 patients classified as receiving palliative-intent ICI therapy confirmed metastatic or unresectable disease in all reviewed cases. Comorbidity data were available for only 63.2% of patients. Finally, tumour-specific analyses were limited by small numbers of vaccine-exposed patients.

In summary, conventional fixed-window analyses produced associations between COVID-19 vaccination and longer overall survival only when vaccination exposure was allowed to extend across the ICI treatment period. For mRNA vaccination, this association became non-significant in calendar-restricted and supportive time-varying analyses, while landmark, initial time-varying, pre-ICI and active vaccinated comparator analyses likewise showed no significant association between mRNA vaccination and overall survival. Together with recent large-scale and methodologically focused studies, these findings suggest that vaccination timing, calendar era, comparator selection and healthy-vaccinee effects can materially influence retrospective survival estimates. Prospective clinical studies are required to determine whether COVID-19 mRNA vaccination at specific time points during ICI therapy translates into improved clinical outcomes and whether any such effect differs by tumour type.

## Acknowledgements

The authors thank Dr Keren Turton and Dr Nicola Thompson for their constructive comments on an early version of the manuscript.

## Contributions

D.M.F designed the study. J.L.K, M.T.K.C, A.J.D, L.A and D.M.F extracted clinical data. D.M.F and M.T.K.C performed data and initial statistical analyses. D.M.F performed the statistical analyses for the revisions. D.M.F wrote the manuscript. D.M.F, M.T.K.C, J.L.K and S.F contributed to editing.

## Ethics approval

Ethical approval was granted by Cambridge University Hospitals NHS Foundation Trust (ID7592; PRN13592).

## Data availability statement

Aggregate data supporting the findings are provided in the article and supplementary material.

## Open access

For the purpose of open access, the MRC Laboratory of Molecular Biology has applied a CC BY public copyright licence to any Author Accepted Manuscript version arising.

## References

Absalyamov, R., Jasper, R. T., Behar, H. V., Tal, R., Dardik, M., Yaniv, N., Eliakim-Raz, N. & Stemmer, S. M. 2026. COVID-19 mRNA Vaccination Status and Survival in Patients With Cancer on Immune Checkpoint Inhibitors. JCO Oncology Advances, 3, e2600032.

Arbel, R., Rokach, L., Razi, T. & Tadmor, T. 2026. Validation of “SARS-CoV-2 mRNA vaccines sensitize tumors to immune checkpoint blockade” in an independent cohort of 4,407 patients. Cancer Lett, 643, 218319.

Cox, D. R. 1972. Regression Models and Life-Tables. Journal of the Royal Statistical Society: Series B (Methodological), 34, 187–202.

Dumas, E., Gougis, P., Gasparollo, L.Spano, J.-P. & Stensrud, M. J. 2026. Time-related biases in observational studies : an illustration with COVID-19 vaccination during immune checkpoint inhibitor therapy. medRxiv, 2025.11.03.25339367.

Grippin, A. J., Marconi, C., Copling, S., Li, N., Braun, C., Woody, C., Young, E., Gupta, P., Wang, M., Wu, A., Jeong, S. D., Soni, D., Weidert, F., Xie, C., Goldenberg, E., Kim, A., Zhao, C., Devries, A., Castillo, P., Lohray, R., Rooney, M. K., Schrank, B. R., Wang, Y., Ma, Y., Chang, E., Kouzy, R., Dyson, K., Jafarnia, J., Nariman, N., Gladish, G., New, J., Argueta, A., Amaya, D., Thomas, N., Doty, A., Chen, J., Copling, N., Alatrash, G., Simon, J., Davies, A. B., Dennis, W., Liang, R., Lewis, J., Wei, X., Rinsurongkawong, W., Vaporciyan, A. A., Johns, A., Aaroe, A., Abraham, S., Andrews, L., Badami, K. K., Baganz, J. A., Bajwa, P., Barbosa, G. R., Beird, H. C., Brock, K., Burton, E. M., Cata, J., Chung, C., Claussen, C., Crommett, J., Cutherell, M., Dabaja, B., Dagher, H., Daniels, K. M., Domask, M., Draetta, G., Edelkamp, P., Fisher, S., French, K. E., Futreal, A., Gaeta, M., Godoy, M., Goldstein, D., Gunther, J., Hutcheson, K., Jaffray, D., Jin, J., John, T. M., Kell, T., Knafl, M., Kwan, R. C., Lee, J. J., Litton, J., Mcenery, K. W., Mcguire, M., Mescher, B., Musunuru, T., Muthu, M., Nates, J., Owen, C. S., Padmakumar, P., Palaskas, N., Patel, J. J., Prabhakaran, S., Ramsey, L., Ravi, V., Hernandez, C. R., Sajith, B., Scheet, P. A., et al. 2025. SARS-CoV-2 mRNA vaccines sensitize tumours to immune checkpoint blockade. Nature, 647, 488–497.

Heudel, P. E., Avrillon, V., Mastroianni, B., Boyle, H., Assaad, S., Ray-Coquard, I., Spano, J. P., Chabaud, S., Cropet, C., Perol, M. & Blay, J. Y. 2026. COVID-19 vaccination before immune checkpoint inhibitor initiation and the overall survival of patients with advanced cancer: A cohort study. Eur J Cancer, 241, 116770.

Jee, J., Zhang, J., Lavery, J. A., Waters, M., Fong, C. J., Minn, A. J., Glickman, M. S., Panageas, K. S., Sawyers, C. L. & Schultz, N. 2026. Deconvolving SARS-CoV-2 mRNA vaccine impact on immunotherapy-related survival. Cancer Discov, 16, 2235–2241.

Jourdain, H., Haddy, N., Singier, A., Duchemin, T., Semenzato, L., Weill, A., Rachas, A., Vignot, S. & Zureik, M. 2026. Overall survival according to COVID-19 mRNA vaccination co-exposure in patients treated with PD-(L)1 inhibitors: a French real-world cohort study. Ann Oncol, 37, 1186–1190.

Karadakic, R., Keating, N. L. & Barnett, M. L. 2026. Interpreting vaccine-associated survival differences in immune checkpoint inhibitor therapy. Proc Natl Acad Sci U S A, 123, e2621501123.

Lin, S. S., Liu, J. F., Hsu, P. S., Lai, H. C. & Cheng, W. P. 2026. Reconciling Conflicting Evidence on Peri-Immune Checkpoint Inhibitor COVID-19 mRNA Vaccination in Lung Cancer: A Time-Aligned Target Trial Emulation-Informed Landmark Study. Value Health.

Salem, J.-E., Curmin, R., Ajrouche, A., De Rycke, Y., Cadranel, J. & Tubach, F. 2026. COVID-19 vaccination around immune checkpoint inhibitor start and survival in a nationwide cohort of patients with cancer. Nature Medicine, 32, 3633–3643.

Therneau, T. M. & Grambsch, P. M. 2000. Modeling Survival Data: Extending the Cox Model. Statistics in the health sciences.

