## Supplementary Data for "Association of mRNA COVID-19 vaccination with overall survival in patients receiving immune checkpoint inhibitors: a retrospective cohort analysis"

**Contents:**

**Page 2: Supplementary Fig. 1.** Schematic overview of vaccination exposure definitions and complementary analytical approaches used in the survival analyses.

**Page 5: Supplementary Table 1.** COVID-19 vaccination status and timing according to the initial exposure window among patients treated with palliative-intent ICI therapy, stratified by tumour types.

**Page 6: Supplementary Table 2**. Association between COVID-19 vaccination within 30 days before ICI initiation and overall survival.

**Page 8: Supplementary Table 3**. COVID-19 vaccination status and timing according to the extended exposure window among patients treated with palliative-intent ICI therapy, stratified by tumour types.

**Page 9: Supplementary Table 4**. Additional sensitivity analyses of ICI initiation year and calendar era for the association between COVID-19 mRNA vaccination and overall survival in patients receiving palliative-intent ICI therapy.

**Page 10: Supplementary Table 5**. Exploratory tumour-specific adjusted analyses of COVID-19 mRNA vaccination and overall survival in patients receiving palliative-intent ICI therapy.

**Page 12: Supplementary Table 6**. Sensitivity analyses incorporating pre-ICI ECOG performance status and comorbidity burden in adjusted analyses of COVID-19 mRNA vaccination and overall survival in patients receiving palliative-intent ICI therapy.

**Page 14: Supplementary Table 7**. Proportional-hazards diagnostics for the principal adjusted Cox models of COVID-19 vaccination and overall survival in patients receiving palliative-intent ICI therapy.

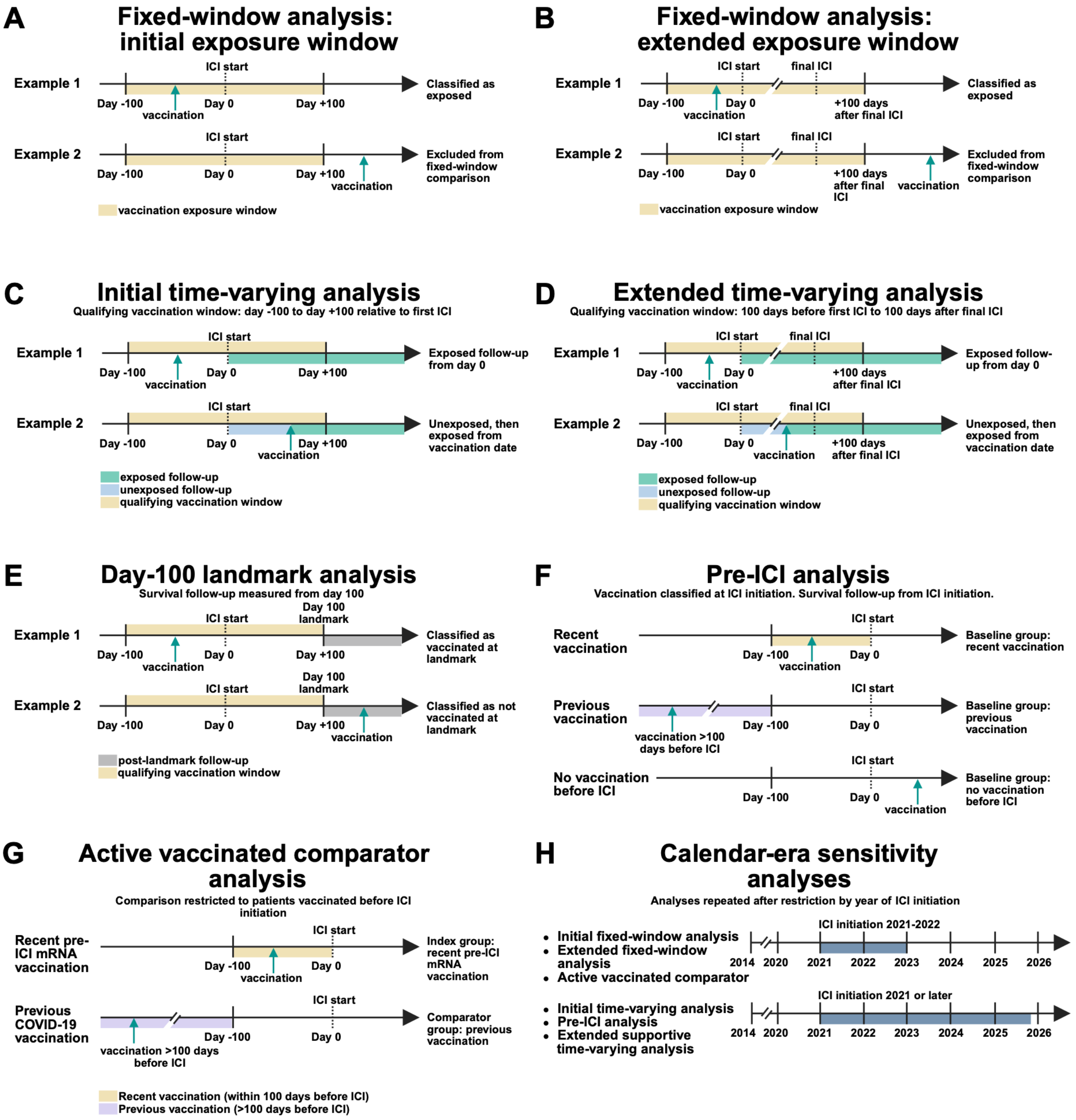


**Supplementary Fig. 1. Schematic overview of vaccination exposure definitions and complementary analytical approaches used in the survival analyses. A**, Initial fixed-window analysis: vaccination exposure was defined by COVID-19 vaccination from 100 days before to 100 days after the first palliative-intent ICI cycle. Survival follow-up began at ICI initiation. **B**, Extended fixed-window analysis: the exposure window extended from 100 days before the first palliative-intent ICI cycle to 100 days after the final palliative-intent ICI cycle. For both fixed-window analyses, patients with no recorded COVID-19 vaccination formed the comparator group, while patients vaccinated exclusively outside the relevant exposure window were excluded from the comparison. **C**, Initial time-varying analysis: qualifying vaccination was restricted to the initial -100 to +100-day window. Patients vaccinated before ICI initiation were considered exposed from day 0. Patients vaccinated after ICI initiation contributed unexposed follow-up until vaccination and exposed follow-up after vaccination. **D**, Extended time-varying analysis: qualifying vaccination extended from 100 days before the first palliative-intent ICI cycle to 100 days after the final palliative-intent ICI cycle. Patients vaccinated before ICI initiation were exposed from day 0, while those vaccinated after ICI initiation contributed unexposed follow-up until vaccination and exposed follow-up after vaccination. This analysis was considered supportive because whether a vaccination qualified depended on the retrospectively determined final ICI cycle. **E**, Day-100 landmark analysis: patients were required to be alive and under observation beyond day 100. Follow-up began at day 100 and vaccination exposure was defined using the initial -100 to +100-day window. Patients vaccinated before the landmark but exclusively outside this window were excluded from the pairwise comparison, while vaccination after day 100 did not alter landmark classification. **F**, Pre-ICI analysis: baseline vaccination status was determined from vaccination history available at ICI initiation. Patients were classified as recently vaccinated (defined as vaccination within 100 days before ICI initiation), previously vaccinated (vaccination more than 100 days before ICI initiation with no vaccination during the preceding 100 days), or having no vaccination before ICI. Vaccination after ICI initiation did not alter baseline classification. **G**, Active vaccinated comparator analysis: patients receiving mRNA vaccination within 100 days before ICI initiation were compared with patients previously vaccinated against COVID-19 but with no vaccination during the preceding 100 days. **H**, Calendar-era sensitivity analyses: initial and extended fixed-window analyses and the active vaccinated comparator analysis were repeated among patients initiating ICI during 2021-2022. Initial time-varying, pre-ICI and extended supportive time-varying analyses were additionally repeated among patients initiating ICI in 2021 or later. In the time-varying analyses, mRNA and AdV vaccination were modelled simultaneously. Abbreviations: AdV - adenoviral-vector; ICI - immune checkpoint inhibitor.

| Tumour group | Total patients treated with palliative-intent ICI, n | At least one COVID-19 vaccination, n | No recorded COVID-19 vaccination, n | Total vaccinated within initial exposure window, n | mRNA-only (within initial exposure window), n | AdV-only (within initial exposure window), n | Mixed (mRNA + AdV within initial exposure window), n | Vaccinated exclusively outside initial exposure window, n |
|---|---|---|---|---|---|---|---|---|
| **All** | 2054 | 493 | 1561 | 148 | 76 | 70 | 2 | 345 |
| **Kidney cancer** | 462 | 138 | 324 | 50 | 22 | 27 | 1 | 88 |
| **Melanoma** | 465 | 127 | 338 | 23 | 11 | 11 | 1 | 104 |
| **NSCLC** | 311 | 58 | 253 | 19 | 13 | 6 | 0 | 39 |
| **Remaining tumour types** | 816 | 170 | 646 | 56 | 30 | 26 | 0 | 114 |
| Head and neck | 149 | 35 | 114 | 13 | 10 | 3 | 0 | 22 |
| Skin SCC | 85 | 9 | 76 | 2 | 1 | 1 | 0 | 7 |
| Gynaecological | 79 | 24 | 55 | 8 | 6 | 2 | 0 | 16 |
| Hepatobiliary | 77 | 19 | 58 | 3 | 1 | 2 | 0 | 16 |
| Mesothelioma (lung) | 68 | 17 | 51 | 9 | 5 | 4 | 0 | 8 |
| Lower GI tract | 55 | 21 | 34 | 8 | 3 | 5 | 0 | 13 |
| Lung small-cell | 53 | 9 | 44 | 4 | 0 | 4 | 0 | 5 |
| Bladder | 40 | 6 | 34 | 2 | 0 | 2 | 0 | 4 |
| Oesophagus | 38 | 7 | 31 | 2 | 0 | 2 | 0 | 5 |
| Breast | 24 | 4 | 20 | 2 | 2 | 0 | 0 | 2 |
| Urothelial | 21 | 4 | 17 | 0 | 0 | 0 | 0 | 4 |
| Gastric | 20 | 2 | 18 | 1 | 0 | 1 | 0 | 1 |
| Sarcoma | 17 | 5 | 12 | 0 | 0 | 0 | 0 | 5 |
| Merkel | 16 | 1 | 15 | 0 | 0 | 0 | 0 | 1 |
| Pancreas | 15 | 1 | 14 | 0 | 0 | 0 | 0 | 1 |
| Cervix | 13 | 1 | 12 | 0 | 0 | 0 | 0 | 1 |
| Prostate | 7 | 1 | 6 | 0 | 0 | 0 | 0 | 1 |
| Haematological | 6 | 1 | 5 | 0 | 0 | 0 | 0 | 1 |
| Skin BCC | 6 | 0 | 6 | 0 | 0 | 0 | 0 | 0 |
| Lung large-cell | 5 | 0 | 5 | 0 | 0 | 0 | 0 | 0 |
| Neuroendocrine | 4 | 1 | 3 | 0 | 0 | 0 | 0 | 1 |
| CUP | 3 | 1 | 2 | 1 | 1 | 0 | 0 | 0 |
| Gallbladder | 3 | 0 | 3 | 0 | 0 | 0 | 0 | 0 |
| Primary brain | 3 | 1 | 2 | 1 | 1 | 0 | 0 | 0 |
| Adrenal | 2 | 0 | 2 | 0 | 0 | 0 | 0 | 0 |
| Mesothelioma (peritoneal) | 2 | 0 | 2 | 0 | 0 | 0 | 0 | 0 |
| Thymic | 2 | 0 | 2 | 0 | 0 | 0 | 0 | 0 |
| NUT carcinoma | 1 | 0 | 1 | 0 | 0 | 0 | 0 | 0 |

| | | | | | | | | |
|---|---|---|---|---|---|---|---|---|
| Skin eccrine adenocarcinoma | 1 | 0 | 1 | 0 | 0 | 0 | 0 | 0 |
| Thyroid | 1 | 0 | 1 | 0 | 0 | 0 | 0 | 0 |

**Supplementary Table 1. COVID-19 vaccination status and timing according to the initial exposure window among patients treated with palliative-intent ICI therapy, stratified by tumour types**. The initial exposure window was defined as receipt of at least one COVID-19 vaccine dose from 100 days before to 100 days after the first palliative-intent ICI cycle. Indented rows report tumour types comprising the 'Remaining tumour types' category. mRNA-only and AdV-only indicate exposure only to that vaccine platform within the window, while mixed indicates exposure to both. Abbreviations: AdV – adenoviral-vector; BCC - basal cell carcinoma; CUP - cancer of unknown primary; GI - gastrointestinal; ICI - immune checkpoint inhibitor; NSCLC - non-small cell lung cancer; SCC - squamous cell carcinoma.

| Vaccine | ICI initiation | Vaccinated ≤ 30 days, n | Unvaccinated, n | Adjusted HR (95% CI) | *p* value |
|---|---|---|---|---|---|
| mRNA | Full cohort | 26 | 1,778 | 0.89 (0.55-1.44) | 0.637 |
| | 2021-2022 | 26 | 295 | 0.93 (0.56-1.55) | 0.787 |
| AdV | Full cohort | 16 | 1,778 | 1.22 (0.69-2.15) | 0.500 |
| | 2021-2022 | 16 | 295 | 1.30 (0.70-2.39) | 0.403 |

**Supplementary Table 2. Association between COVID-19 vaccination within 30 days before ICI initiation and overall survival.** Vaccinated patients received the specified vaccine platform within 30 days before ICI initiation (days -30 to 0). No patients received both vaccine platforms within this window. Patients vaccinated with the relevant vaccine platform more than 30 days before ICI initiation were included as a separate group in the Cox models. Unvaccinated patients had no recorded COVID-19 vaccination on or before ICI initiation. Hazard ratios were adjusted for age, sex, line of systemic therapy and year of ICI initiation, with baseline hazards stratified by tumour group. Abbreviations: AdV - adenoviral-vector; CI - confidence interval; HR - hazard ratio; ICI - immune checkpoint inhibitor.

| Tumour group | Total patients treated with palliative-intent ICI, n | At least one COVID-19 vaccination, n | No recorded COVID-19 vaccination, n | Total vaccinated within extended exposure window, n | mRNA-only (within extended exposure window), n | AdV-only (within extended exposure window), n | Mixed (mRNA + AdV within extended exposure window), n | Vaccinated exclusively outside extended exposure window, n |
|---|---|---|---|---|---|---|---|---|
| **All** | 2054 | 493 | 1561 | 234 | 110 | 105 | 19 | 259 |
| **Kidney cancer** | 462 | 138 | 324 | 73 | 32 | 32 | 9 | 65 |
| **Melanoma** | 465 | 127 | 338 | 50 | 21 | 24 | 5 | 77 |
| **NSCLC** | 311 | 58 | 253 | 31 | 15 | 16 | 0 | 27 |
| **Remaining tumour types** | 816 | 170 | 646 | 80 | 42 | 33 | 5 | 90 |
| Head and neck | 149 | 35 | 114 | 18 | 12 | 6 | 0 | 17 |
| Skin SCC | 85 | 9 | 76 | 2 | 1 | 1 | 0 | 7 |
| Gynaecological | 79 | 24 | 55 | 16 | 9 | 7 | 0 | 8 |
| Hepatobiliary | 77 | 19 | 58 | 4 | 1 | 2 | 1 | 15 |
| Mesothelioma (lung) | 68 | 17 | 51 | 9 | 5 | 4 | 0 | 8 |
| Lower GI tract | 55 | 21 | 34 | 11 | 5 | 4 | 2 | 10 |
| Lung small-cell | 53 | 9 | 44 | 6 | 2 | 3 | 1 | 3 |
| Bladder | 40 | 6 | 34 | 3 | 1 | 1 | 1 | 3 |
| Oesophagus | 38 | 7 | 31 | 4 | 0 | 4 | 0 | 3 |
| Breast | 24 | 4 | 20 | 2 | 2 | 0 | 0 | 2 |
| Urothelial | 21 | 4 | 17 | 0 | 0 | 0 | 0 | 4 |
| Gastric | 20 | 2 | 18 | 2 | 1 | 1 | 0 | 0 |
| Sarcoma | 17 | 5 | 12 | 0 | 0 | 0 | 0 | 5 |
| Merkel | 16 | 1 | 15 | 0 | 0 | 0 | 0 | 1 |
| Pancreas | 15 | 1 | 14 | 0 | 0 | 0 | 0 | 1 |
| Cervix | 13 | 1 | 12 | 0 | 0 | 0 | 0 | 1 |
| Prostate | 7 | 1 | 6 | 1 | 1 | 0 | 0 | 0 |
| Haematological | 6 | 1 | 5 | 0 | 0 | 0 | 0 | 1 |
| Skin BCC | 6 | 0 | 6 | 0 | 0 | 0 | 0 | 0 |
| Lung large-cell | 5 | 0 | 5 | 0 | 0 | 0 | 0 | 0 |
| Neuroendocrine | 4 | 1 | 3 | 0 | 0 | 0 | 0 | 1 |
| CUP | 3 | 1 | 2 | 1 | 1 | 0 | 0 | 0 |
| Gallbladder | 3 | 0 | 3 | 0 | 0 | 0 | 0 | 0 |
| Primary brain | 3 | 1 | 2 | 1 | 1 | 0 | 0 | 0 |
| Adrenal | 2 | 0 | 2 | 0 | 0 | 0 | 0 | 0 |
| Mesothelioma (peritoneal) | 2 | 0 | 2 | 0 | 0 | 0 | 0 | 0 |
| Thymic | 2 | 0 | 2 | 0 | 0 | 0 | 0 | 0 |
| NUT carcinoma | 1 | 0 | 1 | 0 | 0 | 0 | 0 | 0 |
| Skin eccrine adenocarcinoma | 1 | 0 | 1 | 0 | 0 | 0 | 0 | 0 |
| Thyroid | 1 | 0 | 1 | 0 | 0 | 0 | 0 | 0 |

**Supplementary Table 3. COVID-19 vaccination status and timing according to the extended exposure window among patients treated with palliative-intent ICI therapy, stratified by tumour types**. The extended exposure window was defined as receipt of at least one COVID-19 vaccine dose from 100 days before the first palliative-intent ICI cycle to 100 days after the final ICI cycle. Indented rows report tumour types comprising the 'Remaining tumour types' category. mRNA-only and AdV-only indicate exposure only to that vaccine platform within the window, while mixed indicates exposure to both. Abbreviations: AdV - adenoviral-vector; BCC - basal cell carcinoma; CUP - cancer of unknown primary; GI - gastrointestinal; ICI - immune checkpoint inhibitor; NSCLC - non-small cell lung cancer; SCC - squamous cell carcinoma.

| Analysis | ICI initiation year / calendar-era specification | Comparison | n | mRNA-exposed, n | Adjusted HR (95% CI) | *p* value |
|---|---|---|---|---|---|---|
| **Year of ICI initiation modelled categorically** | | | | | | |
| Initial exposure window: fixed-window | Year of ICI initiation modelled categorically | mRNA-only vs no recorded vaccination | 1637 | 76 | 0.94 (0.67-1.32) | 0.717 |
| Initial exposure window: time-varying | Year of ICI initiation modelled categorically | mRNA-exposed vs mRNA-unexposed follow-up | 2054 | 78 | 0.88 (0.64-1.20) | 0.419 |
| Pre-ICI | Year of ICI initiation modelled categorically | mRNA within 100 days before palliative ICI vs no vaccination before ICI | 1833 | 55 | 1.04 (0.73-1.50) | 0.821 |
| Extended exposure window: fixed-window | Year of ICI initiation modelled categorically | mRNA-only vs no recorded vaccination | 1671 | 110 | 0.62 (0.47-0.81) | <0.001 |
| Extended exposure window: supportive time-varying | Year of ICI initiation modelled categorically | mRNA-exposed vs mRNA-unexposed follow-up | 2054 | 129 | 0.84 (0.66-1.09) | 0.192 |
| **Calendar-era restricted analyses** | | | | | | |
| Initial exposure window: time-varying | ICI initiation 2021 or later | mRNA-exposed vs mRNA-unexposed follow-up | 1321 | 76 | 0.84 (0.62-1.14) | 0.273 |
| Pre-ICI | ICI initiation 2021 or later | mRNA within 100 days before palliative ICI vs no vaccination before ICI | 1100 | 55 | 1.00 (0.70-1.43) | 0.988 |
| Extended exposure window: supportive time-varying | ICI initiation 2021 or later | mRNA-exposed vs mRNA-unexposed follow-up | 1321 | 88 | 0.87 (0.66-1.14) | 0.306 |

**Supplementary Table 4. Additional sensitivity analyses of ICI initiation year and calendar era for the association between COVID-19 mRNA vaccination and overall survival in patients receiving palliative-intent ICI therapy.** Categorical ICI initiation year models included the covariates specified for the adjusted model in the Methods, with calendar year of ICI initiation modelled categorically rather than continuously. Calendar-era analyses were restricted to patients initiating palliative-intent ICI therapy as indicated. For time-varying analyses, the reported mRNA estimates were derived from models that also included AdV vaccination. Abbreviations: AdV - adenoviral-vector; CI - confidence interval; HR - hazard ratio; ICI - immune checkpoint inhibitor.

| Analysis | Tumour group | Comparison | n | mRNA-exposed, n | Adjusted HR (95% CI) | *p* value |
|---|---|---|---|---|---|---|
| Initial exposure window: day-100 landmark | Kidney cancer | mRNA vs not vaccinated at day 100 | 357 | 22 | 0.98 (0.55-1.73) | 0.938 |
| | Melanoma | mRNA vs not vaccinated at day 100 | 365 | 11 | 1.21 (0.53-2.78) | 0.648 |
| | NSCLC | mRNA vs not vaccinated at day 100 | 250 | 12 | 1.80 (0.97-3.34) | 0.061 |
| | Remaining tumour types | mRNA vs not vaccinated at day 100 | 583 | 28 | 0.89 (0.56-1.41) | 0.610 |
| Initial exposure window: time-varying | Kidney cancer | mRNA-exposed vs mRNA-unexposed follow-up | 462 | 23 | 0.76 (0.45-1.29) | 0.303 |
| | Melanoma | mRNA-exposed vs mRNA-unexposed follow-up | 465 | 12 | 0.86 (0.39-1.90) | 0.702 |
| | NSCLC | mRNA-exposed vs mRNA-unexposed follow-up | 311 | 13 | 1.60 (0.94-2.73) | 0.084 |
| | Remaining tumour types | mRNA-exposed vs mRNA-unexposed follow-up | 816 | 30 | 0.79 (0.51-1.22) | 0.294 |
| Pre-ICI | Kidney cancer | mRNA within 100 days before palliative ICI vs no vaccination before ICI | 405 | 15 | 0.63 (0.30-1.35) | 0.239 |
| | Melanoma | mRNA within 100 days before palliative ICI vs no vaccination before ICI | 428 | 8 | 0.85 (0.31-2.31) | 0.753 |
| | NSCLC | mRNA within 100 days before palliative ICI vs no vaccination before ICI | 285 | 11 | 1.46 (0.77-2.77) | 0.244 |
| | Remaining tumour types | mRNA within 100 days before palliative ICI vs no vaccination before ICI | 715 | 21 | 1.08 (0.66-1.75) | 0.767 |
| Extended exposure window: supportive time-varying | Kidney cancer | mRNA-exposed vs mRNA-unexposed follow-up | 462 | 41 | 0.67 (0.44-1.02) | 0.064 |
| | Melanoma | mRNA-exposed vs mRNA-unexposed follow-up | 465 | 26 | 0.71 (0.35-1.44) | 0.342 |
| | NSCLC | mRNA-exposed vs mRNA-unexposed follow-up | 311 | 15 | 1.49 (0.88-2.52) | 0.143 |
| | Remaining tumour types | mRNA-exposed vs mRNA-unexposed follow-up | 816 | 47 | 0.83 (0.58-1.19) | 0.305 |

**Supplementary Table 5. Exploratory tumour-specific adjusted analyses of COVID-19 mRNA vaccination and overall survival in patients receiving palliative-intent ICI therapy.** Within each tumour group, models were adjusted for age, sex, year of ICI initiation and line of systemic therapy at palliative ICI initiation (categorised as first-line versus second-line or later). For time-varying analyses, the reported mRNA estimates were derived from models that also included AdV vaccination. Tumour-specific analyses were exploratory and were not adjusted for multiple comparisons. Abbreviations: AdV - adenoviral-vector; CI - confidence interval; HR - hazard ratio; ICI - immune checkpoint inhibitor; NSCLC - non-small cell lung cancer.

| Analysis | Sensitivity model | Comparison | n | mRNA-exposed, n | Adjusted HR (95% CI) | *p* value |
|---|---|---|---|---|---|---|
| **ECOG sensitivity analyses** | | | | | | |
| Initial exposure window - fixed-window | ECOG-adjusted model | mRNA-only vs no recorded vaccination | 1629 | 76 | 0.81 (0.61-1.08) | 0.149 |
| Initial exposure window - day-100 landmark | ECOG-adjusted model | mRNA vs not vaccinated at day 100 | 1547 | 73 | 1.04 (0.78-1.39) | 0.785 |
| Initial exposure window - time-varying | ECOG-adjusted model | mRNA-exposed vs mRNA-unexposed follow-up | 2044 | 78 | 0.89 (0.68-1.17) | 0.408 |
| Pre-ICI | ECOG-adjusted model | mRNA within 100 days before palliative ICI vs no vaccination before ICI | 1823 | 55 | 1.00 (0.72-1.39) | 0.983 |
| Extended exposure window - fixed-window | ECOG-adjusted model | mRNA-only vs no recorded vaccination | 1663 | 110 | 0.59 (0.46-0.76) | <0.001 |
| Extended exposure window - supportive time-varying | ECOG-adjusted model | mRNA-exposed vs mRNA-unexposed follow-up | 2044 | 129 | 0.83 (0.66-1.04) | 0.105 |
| **Comorbidity sensitivity analyses** | | | | | | |
| Initial exposure window: day-100 landmark | Comorbidity-adjusted model | mRNA vs not vaccinated at day 100 | 986 | 58 | 1.16 (0.84-1.60) | 0.363 |
| Initial exposure window: time-varying | Comorbidity-adjusted model | mRNA-exposed vs mRNA-unexposed follow-up | 1298 | 62 | 0.96 (0.71-1.31) | 0.817 |
| Pre-ICI | Comorbidity-adjusted model | mRNA within 100 days before palliative ICI vs no vaccination before ICI | 1140 | 45 | 1.03 (0.71-1.48) | 0.887 |
| Extended exposure window: supportive time-varying | Comorbidity-adjusted model | mRNA-exposed vs mRNA-unexposed follow-up | 1298 | 92 | 0.96 (0.73-1.25) | 0.740 |

**Supplementary Table 6. Sensitivity analyses incorporating pre-ICI ECOG performance status and comorbidity burden in adjusted analyses of COVID-19 mRNA vaccination and overall survival in patients receiving palliative-intent ICI therapy.** ECOG-adjusted models included the covariates specified for the adjusted model in the Methods, with pre-ICI ECOG performance status added as an additional covariate. ECOG performance status was categorised as 0, 1 or ≥2. Comorbidity-adjusted analyses were restricted to patients with available recorded non-cancer comorbidity data. Of 2,054 patients, 1,298 (63.2%) had recorded comorbidity data. Across these patients, 14 distinct types of non-cancer comorbidity were identified. The number recorded per patient ranged from 0 to 2 comorbidities and was included as a categorical covariate (0, 1 or 2). Of the 1,298 patients with recorded non-cancer comorbidity data, 259 patients had no recorded comorbidities (19.9%), 904 had one (69.7%), and 135 had two (10.4%). The remaining 756 patients (36.8%) had no recorded comorbidity data and were treated as missing data rather than being assigned a zero value. Abbreviations: CI - confidence interval; ECOG - Eastern Cooperative Oncology Group; HR - hazard ratio; ICI - immune checkpoint inhibitor.

| Analysis | Comparison | Exposure-term PH test, $p$ value | Global PH test, $p$ value | Other covariates with evidence of non-proportional hazards |
|---|---|---|---|---|
| **Initial exposure window** | | | | |
| Fixed-window | mRNA-only vs no recorded vaccination | 0.093 | 0.375 | None |
| Fixed-window | AdV-only vs no recorded vaccination | 0.067 | 0.324 | None |
| Day-100 landmark | mRNA vs not vaccinated at day 100 | 0.750 | 0.520 | None |
| Time-varying | mRNA and AdV exposure modelled simultaneously | mRNA: 0.175<br>AdV: 0.237 | 0.102 | Year of ICI initiation: 0.007 |
| Fixed-window, ICI 2021-2022 | mRNA-only vs no recorded vaccination | 0.078 | 0.162 | None |
| **Pre-ICI analyses** | | | | |
| Pre-ICI | mRNA within 100 days before palliative ICI vs no vaccination before ICI | 0.261 | 0.149 | Year of ICI initiation: 0.015 |
| Active vaccinated comparator | mRNA within 100 days before palliative ICI vs previous vaccination >100 days before ICI | 0.099 | 0.033 | None |
| Active vaccinated comparator, ICI 2021-2022 | mRNA within 100 days before palliative ICI vs previous vaccination >100 days before ICI | 0.043 | 0.050 | None |
| **Extended exposure window** | | | | |
| Fixed-window | mRNA-only vs no recorded vaccination | 0.058 | 0.223 | None |
| Fixed-window | AdV-only vs no recorded vaccination | <0.001 | 0.035 | None |
| Fixed-window, ICI 2021-2022 | mRNA-only vs no recorded vaccination | 0.046 | 0.106 | None |
| Supportive time-varying | mRNA and AdV exposure modelled simultaneously | mRNA: 0.330<br>AdV: 0.083 | 0.068 | Year of ICI initiation: 0.006 |

**Supplementary Table 7. Proportional-hazards diagnostics for the principal adjusted Cox models of COVID-19 vaccination and overall survival in patients receiving palliative-intent ICI therapy.** Proportional-hazards (PH) assumptions were assessed using Schoenfeld residual tests. The exposure test evaluated the vaccination exposure term and the global test evaluated the model as a whole. A $p$ value <0.05 indicated evidence against the PH assumption. For an individual model term, this suggested that the corresponding hazard ratio did not remain constant over time. For time-varying analyses, mRNA and AdV vaccination were modelled simultaneously and their exposure-term tests are reported separately. The column ‘other covariates with evidence of non-proportional hazards’ identifies adjustment covariates, other than the vaccination variables, with a PH test $p<0.05$. ‘None’ indicates that no other adjustment covariate showed evidence against the PH assumption. Abbreviations: AdV - adenoviral-vector; ICI - immune checkpoint inhibitor; PH - proportional hazards.
